# The potential contribution of vaccination uptake to occupational differences in risk of SARS-CoV-2: Analysis of the ONS COVID-19 Infection Survey

**DOI:** 10.1101/2023.03.24.23287700

**Authors:** Jack Wilkinson, Evangelia Demou, Mark Cherrie, Rhiannon Edge, Matthew Gittins, S Vittal Katikireddi, Theocharis Kromydas, William Mueller, Neil Pearce, Martie van Tongeren, Sarah Rhodes

**Author notes:** <u>Corresponding author:</u> Dr Jack Wilkinson, Centre for Biostatistics, University of Manchester.

## Abstract

**Objectives:** To assess variation in vaccination uptake across occupational groups as a potential explanation for variation in risk of SARS-CoV-2 infection.

**Design:** We analysed data from the UK Office of National Statistics COVID-19 Infection Survey linked to vaccination data from the National Immunisation Management System in England from December 1^st^ 2020 to 11^th^ May 2022. We analysed vaccination uptake and SARS-CoV-2 infection risk by occupational group and assessed whether adjustment for vaccination reduced the variation in risk between occupational groups.

**Setting:** 

**Results:** Estimated rates of triple-vaccination were high across all occupational groups (80% or above), but were lowest for food processing (80%), personal care (82%), hospitality (83%), manual occupations (84%), and retail (85%). High rates were observed for individuals working in health (95% for office-based, 92% for those in patient-facing roles) and education (91%) and office-based workers not included in other categories (90%). The impact of adjusting for vaccination when estimating relative risks of infection was generally modest (ratio of hazard ratios reduced from 1.38 to 1.32), but was consistent with the hypothesis that low vaccination rates contribute to elevated risk in some groups. Conversely, estimated relative risk for some occupational groups, such as people working in education, remained high despite high vaccine coverage.

**Conclusions:** Variation in vaccination coverage might account for a modest proportion of occupational differences in infection risk. Vaccination rates were uniformly very high in this cohort, which may suggest that the participants are not representative of the general population. Accordingly, these results should be considered tentative pending the accumulation of additional evidence.

## Background

SARS-CoV-2 infection and COVID-19 mortality risks in the United Kingdom have been reported to differ across occupational groups, with variation seen over time (1-5). Some of this variation appears to be driven by workplace factors, such as the number of people in the workplace, ability to socially distance from others, and whether the work is located in an indoor or outdoor environment (6-8). However, uncertainty remains regarding the extent to which occupational differences are driven by these characteristics compared to non-workplace factors.

One plausible contributor to the observed variation in risk may be occupational differences in rates of vaccination uptake against COVID-19. Vaccination appears to reduce the risk of SARS-CoV-2 infection as well as associated morbidity and mortality (9, 10), although effects appear to strongly depend on the time since last vaccination (11). In the UK, vaccine delivery was not mandated but was initially prioritised for certain high-risk occupational groups, such as healthcare workers. Identifying groups with low vaccination is important for informing future vaccine delivery strategy, as it may be necessary to offer specific, targeted encouragement to improve uptake. This would be particularly important if there are groups where low vaccination rates contribute to a higher risk of SARS-CoV-2 transmission in the workplace. Recent studies have highlighted considerable variation in vaccine uptake across occupations (12-14), with low levels observed in people working in elementary trades. However, different data sources have different limitations, such as the possibility of non-random missing data (14). It is therefore crucial to investigate the replicability of findings relating to vaccination uptake across a variety of data sources, to permit triangulation of results (15). Moreover, estimating the variation in risk of SARS-CoV-2 infection across occupations with and without adjustment for vaccination status might indicate whether vaccination uptake is an important factor explaining these occupational differences. We therefore analysed data from the Office for National Statistics (ONS) Coronavirus Infection Survey (CIS) to characterise vaccine uptake across occupational groups and to examine vaccination uptake as a possible explanation for occupational variations in SARS-CoV-2 infection.

## Methods

### Datasets

The CIS has been described elsewhere (2, 16). It is a randomly sampled panel survey of households, including participants aged two years and older, that aims to be representative of the UK population. It began recruitment in April 2020 and has added new participants monthly until January 2022(17). Participants were visited weekly for the five weeks following their recruitment, and monthly thereafter. Survey responses were collected at in-person visits until April 2022, with each visit incorporating a PCR test for SARS-CoV-2, to enable estimation of prevalence. From April 2022 onwards the CIS phased out in-person visits and started using a more flexible data collection approach using online, telephone and postal methods. Recruitment rates were initially high (51%) but eventually dropped to around 12% (18). Rate of attrition was very low (less than 1% in 2021, (18)).

For participants in England, vaccination information (doses received, dates received, vaccine type) is available from the National Immunisation Management System (NIMS)(19). ONS supplied a dataset containing NIMS data, which was bolstered using self-report data from CIS. Because NIMS does not cover Scotland, Wales, and Northern Ireland, only self-report data are available for these countries. This represented a fundamental difference in measurement of vaccination in England compared to the other countries, as any vaccinations taking place after an individual’s most recent CIS response could be accounted for in England, but not for the other countries. This would lead to considerable undercounting outside of England. We therefore restricted analyses to participants resident in England. We used data (from CIS and NIMS) from 1st December 2020, roughly coinciding with the start of the vaccination programme in the UK, and included all survey visits up to 11th May 2022, which was the most recent available data at the time of the analysis.

### Inclusion criteria

Analyses were conducted on working-age participants in England, which we defined as being aged 20-64 years at their first CIS visit. The definition was selected to allow direct comparison with previous studies (2).

### Classification of occupation

Occupational groupings were derived using 4-digit standard occupational codes (SOC). We used a bespoke categorisation system to allow us to distinguish occupations likely to be at high risk of SARS-CoV-2 infection, informed by our previous work on this topic (Supplementary Table 1). We defined the following occupational categories: education; food processing; healthcare (office-based); healthcare (patient contact); hospitality; manual; other workers (non-office-based); other workers (office-based); personal care; police and protective services; retail; sanitation services; social care; transport (non-public-facing); transport (public-facing); not working/ student. Rather than exclude individuals with missing occupation data, we created a ‘missing’ category, distinct from ‘not employed’. The classification scheme is presented in Supplementary Table 1.

**Table 1.** Characteristics of the sample. Mean (SD) or n (%)

| Characteristic | Summary (n = 256,598) |
| --- | --- |
| Age (years) | 45.3 (12.3) |
| Sex male | 115,904 (45) |
| Ethnicity |  |
| White | 231,868 (90) |
| Asian | 13,973 (5) |
| Black | 3,465 (1) |
| Mixed | 4,298 (2) |
| Other | 2,982 (1) |
| Missing | 12 (0) |
| Occupation |  |
| Education | 12,131 (5) |
| Food processing | 1,080 (0) |
| Healthcare-office based | 777 (0) |
| Healthcare patient contact | 10,970 (4) |
| Hospitality | 2,462 (1) |
| Manual | 10,387 (4) |
| Other workers non-office based | 11,351 (4) |
| Other workers office based | 70,210 (27) |
| Personal care | 732 (0) |
| Police and protective services | 3,076 (1) |
| Retail | 5,875 (2) |
| Sanitation services | 1,610 (1) |
| Social care | 6,533 (3) |
| Transport non-public facing | 2,990 (1) |
| Transport public facing | 1,088 (0) |
| Student/ not working | 65,236 (25) |
| Missing/ incomplete | 50,090 (20) |
| IMD |  |
| 1 | 40,810 (16) |
| 2 | 62,239 (24) |
| 3 | 73,118 (29) |
| 4 | 80,431 (31) |
| Urban-rural |  |
| Major urban | 106,046 (41) |
| Urban city town | 105,716 (41) |
| Rural town | 23,318 (9) |
| Rural village | 21,518 (8) |
| Household size |  |
| 1 | 34,764 (14) |
| 2 | 97,856 (38) |
| 3 | 52,013 (20) |
| 4 | 50,666 (20) |
| 5+ | 21,299 (8) |
| Health conditions |  |
| Yes | 44,599 (17) |
| Missing | 3,084 (1) |

### Outcome definition

The outcome variable was SARS-CoV-2 infection, defined as a positive PCR test obtained from a CIS visit. Individuals infected with SARS-CoV-2 may test positive on PCR for an extended duration, including on consecutive CIS visits. This makes it difficult to distinguish repeated positive tests due to a single infection from repeated infections. We dealt with this issue by sensitivity analysis. Specifically, we conducted the analysis using two different definitions for a repeat infection. In the first, any new positive PCR test was treated as a new infection, provided that there was at least one negative PCR test since the previous positive test. In the second, we required a gap of 6 months between positive PCR tests, with at least one negative PCR test in between, to consider a subsequent positive PCR test to represent a new infection. This latter criterion was in line with UK government guidance concerning immunity following infection(20). If results did not substantially differ between these two scenarios, it would indicate that findings were robust to handling of this issue.

### Statistical analysis

We created descriptive summaries of participant characteristics. We summarised vaccination details (number of doses received, type of each dose) by occupation group. We then conducted analyses looking at how risk of SARS-CoV-2 infection varied across occupational groups in the period 1st December 2020 to 11th May 2022, and how adjustment for vaccination status affected these estimates.

We used time-to-event methods, with calendar time as the timescale for analysis. Specifically, we set 1st December 2020 as the time origin, and considered participants to be left-censored prior to their first survey visit after this date. We incorporated multiple infections per individual using a Prentice, Williams, Peterson Total Time approach, which estimates hazard ratios with stratification by event number (21, 22). We included COVID-19 vaccination status as a categorical variable, measuring the number of vaccines received (0,1,2,3). The count of vaccines for each individual was not updated until 14 days after the vaccination date, to allow for a delay in conferred protection from the vaccine. The longitudinal nature of the survey means that repeated measurements are available for participants over time. We allowed all variables to be time-varying in the analysis. These analyses assume that the adjustment set, described below, was sufficient to account for any confounding of the occupational-infection relationship and of the vaccination-infection relationship.

We fitted models sequentially, with the adjustment variables selected using a previously-derived DAG (an interactive version is available at http://dagitty.net/dags.html?id=5J_TeK presented in (2). The DAG was constructed with the aim of identifying short-term effects of attending the workplace, rather than the effects of extended tenure in an occupation. Accordingly, we considered variables relating to health and living conditions to be confounders, rather than mediators, of the occupation effect. We first fitted models with occupation category as the exposure variable, adjusted for age and sex (Model 1). We then additionally adjusted for ethnic group, Index of Multiple Deprivation (IMD, categorised by quartiles), geographic region, household size, urban or rural location and presence of a health condition (Model 2). We then added vaccination status, as described above (Model 3). We present hazard ratios (95% CIs) for each adjustment set. We calculated the ‘ratio of hazard ratios’ as the ratio of the We used ‘other office-based workers’ as the reference category for the occupation variable, as this was considered to be a large, low-risk group. We also present hazard ratios (95% CIs) corresponding to number of vaccines received.

## Results

Our analyses included 256,598 working age adults. Characteristics of the sample are shown in Table 1.

### Number of vaccines by occupation

Figure 1 shows the proportion of survey participants in each occupation category who had zero, one, two, or three (or more) vaccines by May 11^th^ 2022. The overwhelming majority of individuals in all categories had had three vaccines. The proportion of individuals who had had three vaccines was lowest for food processing. Results for office-based healthcare workers, personal care workers, and public-facing transport workers cannot be fully displayed due to disclosure rules. However, office-based healthcare workers had the highest proportion with triple-vaccination (95%). Proportions of individuals who had received three vaccinations were 80% in personal care and 87% in public-facing transport roles.

**Figure 1:**
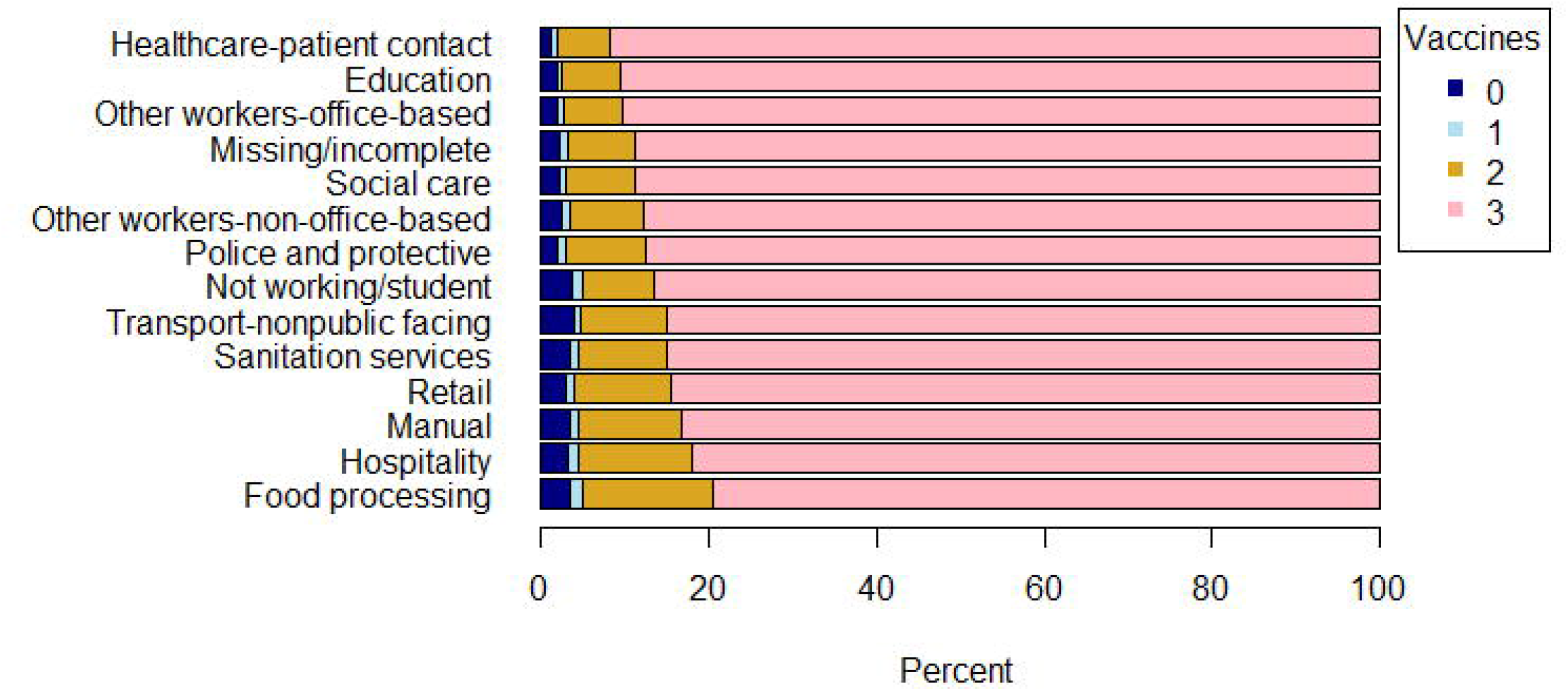
Percentage of individuals receiving 0,1,2,3 COVID-19 vaccines in England by occupation group by May 11^th^ 2022. Note that disclosure rules mean that results for office-based healthcare workers, personal care workers, and public-facing transport workers cannot be displayed.

Supplementary Tables 2, 3, and 4 show the type of vaccine received for first, second, and third doses by occupation group.

### Relationship with infection risk

Supplementary Table 5 shows the estimated hazard ratios for number of vaccines received in relation to SARS-CoV-2 infection. Infection risk decreased with increasing numbers of vaccinations. Supplementary Table 6 shows the number of infections by occupational group, although this is heavily redacted due to disclosure rules. Figure 2 shows risk of infection (at least 1) and proportion of individuals with three vaccinations by occupational group. Risk of infection in this period was not highly variable across groups. Individuals working in personal care and food processing had relatively low rates of triple-vaccination and relatively high rates of infection. People working in education had the highest risk of infection, and high rates of triple-vaccination. People working in healthcare had the highest rates of triple-vaccination, whereas risk of infection was not notably elevated compared to other occupational groups, including those who had regular contact with patients.

**Figure 2:**
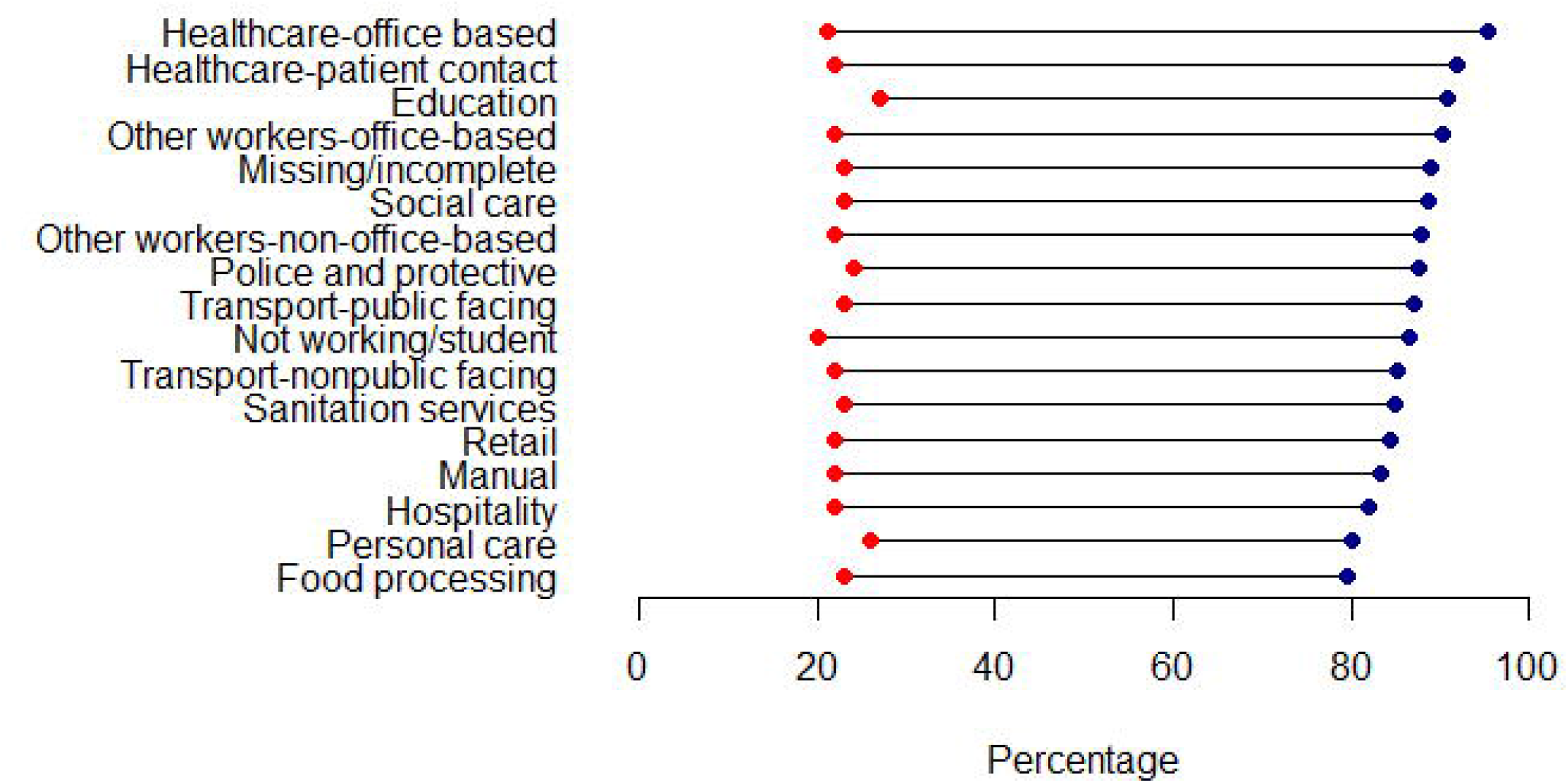
Rates of SARS-CoV-2 infection (at least one, red) and triple vaccination (blue) by occupation, between December 1^st^ 2020 and May 11^th^ 2022.

Estimated hazard ratios (95% CIs) from PWP models are shown in Figure 3 and Supplementary Table 7. Risk of infection during this period remained relatively high for education workers, people working in hospitality, personal care, police and protective services, social care, and potentially public-facing transport roles, compared to other office-based workers. Adjustment for non-vaccine covariates had little impact on estimates. The impact of adjustment for number of vaccines received was negligible for some groups, such as education, social care, and police and protective services. It was more pronounced for others; estimates for individuals employed in food processing, hospitality, manual professions, personal care, retail, sanitation services, and non-public facing transport roles were all noticeably reduced after adjustment for vaccination, albeit with considerable imprecision in estimates. Adjusting for vaccination substantially increased the estimate for healthcare roles with patient-contact, and to a lesser extent office-based healthcare workers. The ratio of hazard ratios reduced from 1.38 prior to adjustment for vaccination (Model 2) to 1.32 following adjustment (Model 3).

**Figure 3:**
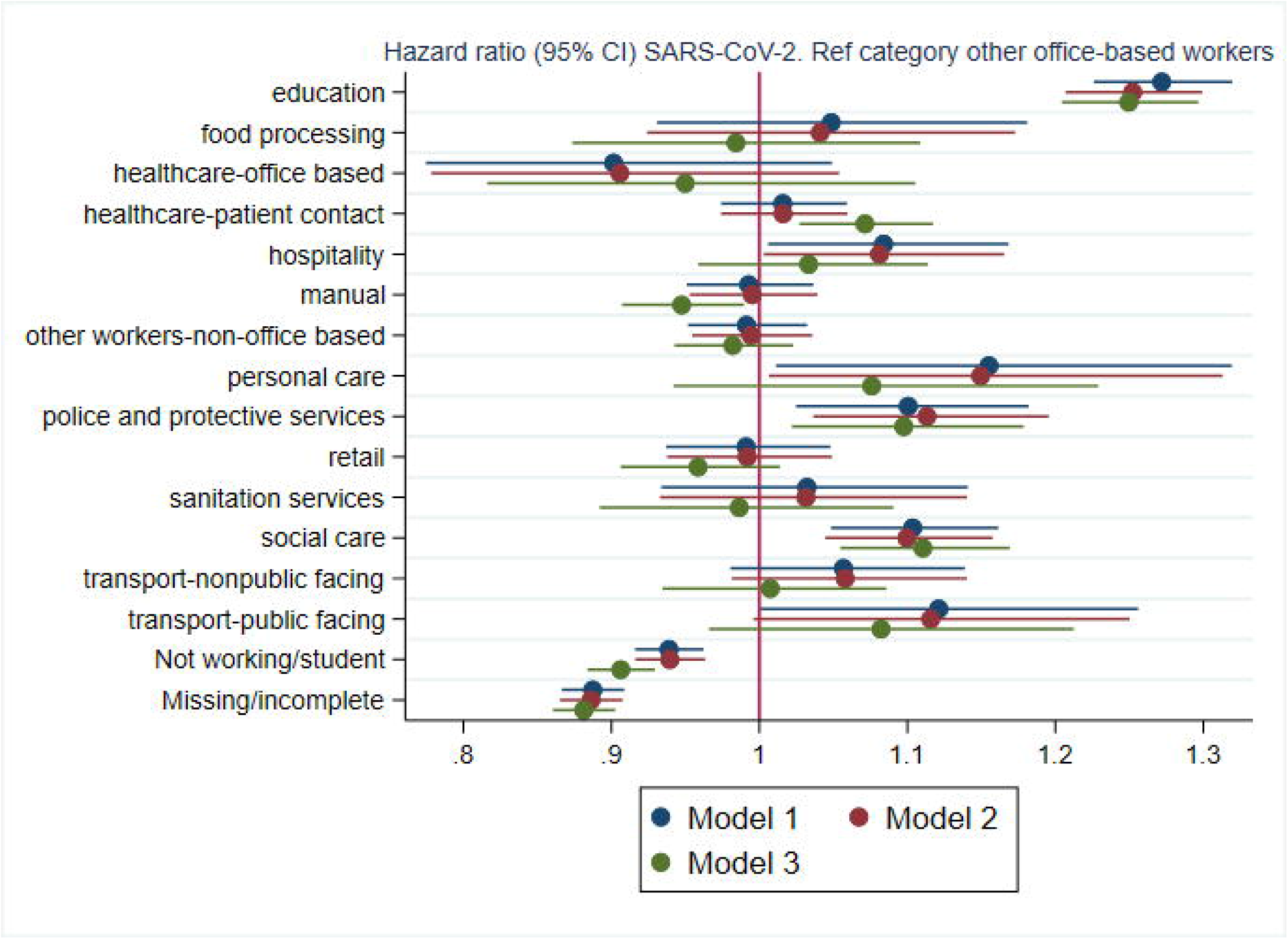
Hazard ratios (95% CIs) corresponding to occupational group in relation to infection with SARS-CoV-2. Based on n = 256,598 individuals. Model 1: Adjusted for age and sex. Model 2: Additionally adjusted for ethnicity, deprivation, region, urban or rural area, household size, and presence of pre-existing health conditions. Model 3: Additionally adjusted for number of vaccines received. The reference category was ‘other office-based workers’.

Results from the sensitivity analysis where a 6-month gap between positive PCR tests was required to register a new infection demonstrated a negligible difference in results (Supplementary Figure 1 and Supplementary Table 8).

## Discussion

The current results suggest that variation in vaccine uptake explains some of the differences in risk of SARS-CoV-2 infection between occupational groups. While the overwhelming majority of individuals in all groups had received three vaccines by 11^th^ May 2022, there was substantial variation in the number of vaccines received between occupations. The variation in absolute risk of infection between occupational groups during the study period was not large.

These results are consistent with the findings from the Virus Watch cohort which found that individuals working in transport, trade, service and sales had the lowest uptake (12), and also with an analysis based on 2021 Census data which suggested high uptake amongst office-based and professional workers and low uptake amongst people working in elementary occupations (14).

The estimates of relative infection risk for occupational groups with the lowest rates of triple-vaccination (food processing, hospitality, manual, personal care, retail, and sanitation services) were all reduced after adjusting for vaccination, which might suggest that relatively low vaccination rates account for some of the risk in these groups. Conversely, relative risk estimates for the occupational groups with the highest rates of triple-vaccination were increased after adjustment for vaccination, suggesting that high vaccination coverage in healthcare workers may have conferred notable protection in this (high risk) group during the study period. However, there were also occupational groups with high vaccination coverage for which relative risk estimates were essentially unaffected after adjusting for vaccination (education, social care). Relative risk of infection remained high for these groups despite vaccination (although for social care, overall rates of triple-vaccination in the period was below that observed for the reference group, other office-based workers). The estimated relative risk for police and protective services remained similar after adjustment for vaccination, and was elevated. Again, vaccination rates in this group were lower than for the reference group.

This is the first study to attempt to reconcile the effect of covid-19 vaccination on the occupational risk of SARS-CoV-2 infection. There are a number of important limitations to consider in interpreting the results. Our analyses assume that we have sufficiently controlled for confounding between occupation and infection, and between vaccination and infection. If the latter condition was not met, adjusting for vaccination as a purported mediator of the occupation-infection relationship could induce collider stratification bias. One probable source of unmeasured confounding in the current study relates to individual behaviour, such as socialising and shopping. However, previous analyses suggest that whilst occupational differentials in SARS-CoV-2 infection risk vary over time, they are relatively robust to adjustment for socio-demographic, health-related, and non-workplace activity-related potential confounders (2, 5). While data on behaviour is captured within the CIS, it is self-reported and subject to substantial rates of missingness. Further investigation of this dataset using methods for time-varying confounding (23, 24) could prove worthwhile.

Another limitation of the dataset is that the relative timing of vaccination and infection is not always clear. If a participant obtains a positive PCR test at a survey visit, we can say that they became infected sometime since the previous visit, but whether this occurred prior to or following vaccination cannot be discerned, since we cannot know the actual date of infection. Another potential source of bias relates to the representativeness of the CIS. Estimated vaccine uptake reported here is similar to that observed in the Virus Watch cohort (12) but is considerably higher compared to some administrative databases, potentially indicating that response rate might be higher in vaccinated individuals. For example, we estimated the proportion of manual workers who had received three vaccinations to be over 80% by 11^th^ May 2022. By contrast, an analysis based on 2021 Census data estimated the proportion of workers in elementary and related occupations to be just 58% by 28^th^ February 2022 (14). It is unclear however if the observed differences are due to lack of representativeness in the CIS, due to biases in the Census-based study arising from missing data, or due to other methodological differences, such as the discrepancy in study periods. Overall, 88% of the CIS cohort had received three vaccinations by May 11^th^ 2022. For comparison, we estimate that approximately 64% of 18-64 year olds in England had had three vaccinations by this date using publically-available NIMS (25)and Census 2011 (26) data. Noting the slight difference in age bandings, it does therefore appear that vaccination rates may be higher in the CIS cohort. It may be, for example, that participation in the study increases the likelihood of vaccination. If vaccination rates are consistently overestimated in the CIS data, this could cause us to understate the role of vaccination in explaining differential risk between occupations, and could plausibly mean that overall variation in risk between occupations is understated. Consequently, triangulation of the results relating to vaccination effects will be important, using data sources with different biases, although at present there is no comparable analysis examining the role of vaccination in explaining differences in infection risk. These results should be considered preliminary pending the accrual of further relevant data. We did find that vaccinations rates were lowest amongst several occupational groups involving substantial public contact, in line with recent work (14).

## Conclusions

The present results suggest that differences in vaccination uptake between occupations contribute to some of the difference in infection risk. However, it is not sufficient to explain all of the variation in risk, and important differences remain. These could be related to workplace factors, work activities, or behaviours outside the workplace. Complementary approaches are therefore likely to remain necessary, particularly for high-risk occupations.

## Data Availability

ONS CIS data can be accessed only by researchers who are Office of National Statistics (ONS) accredited researchers. Researchers can apply for accreditation through the Research Accreditation Service. Access is through the Secure Research Service (SRS) and approved on a project basis. For further details see: https://www.ons.gov.uk/aboutus/whatwedo/statistics/requestingstatistics/approvedresearcherscheme

## Contributors

SR is principal investigator. All authors contributed to the design of the research proposal and study, including the statistical analysis plan. JW conducted analyses and wrote first draft of the manuscript. All authors contributed to the interpretation of the results, critically revised the paper and agreed on the final version for submission.

## Disclaimer

This work contains statistical data from ONS which is Crown Copyright. The use of the ONS statistical data in this work does not imply the endorsement of the ONS in relation to the interpretation or analysis of the statistical data. This work uses research datasets which may not exactly reproduce National Statistics aggregates.

## Ethics approval

The COVID-19 Infection Survey received ethical approval from the South-Central Berkshire B Research Ethics Committee (20/SC/0195). All participants provided informed consent. For use of this data for this project statistics authority self-assessment classified the study as low risk. This assessment was approved by the Office for National Statistics (ONS) Research Accreditation Panel.

## Patient and public involvement

Participants were not involved in the design and implementation of the study or in setting research questions and the outcome measures. No participants were asked to advise on interpretation or writing up of results.

## Acknowledgements

We would like to acknowledge Vahé Nafilyan, Office for National Statistics, for substantial input into the contents of this manuscript.

