## Supplementary for "The potential contribution of vaccination uptake to occupational differences in risk of SARS-CoV-2: Analysis of the ONS COVID-19 Infection Survey"

**Supplementary Tables**

| SOC2010 | NEW OCCUPATIONAL GROUPINGS | Occ_subgroups |
| --- | --- | --- |
| 1115 | Chief executives and senior officials | other workers-office based |
| 1116 | Elected officers and representatives | other workers-office based |
| 1121 | Production managers and directors in manufacturing | other workers-office based |
| 1122 | Production managers and directors in construction | other workers-office based |
| 1123 | Production managers and directors in mining and energy | other workers-office based |
| 1131 | Financial managers and directors | other workers-office based |
| 1132 | Marketing and sales directors | other workers-office based |
| 1133 | Purchasing managers and directors | other workers-office based |
| 1134 | Advertising and public relations directors | other workers-office based |
| 1135 | Human resource managers and directors | other workers-office based |
| 1136 | Information technology and telecommunications directors | other workers-office based |
| 1139 | Functional managers and directors n.e.c. | other workers-office based |
| 1150 | Financial institution managers and directors | other workers-office based |
| 1161 | Managers and directors in transport and distribution | other workers-office based |
| 1162 | Managers and directors in storage and warehousing | other workers-office based |
| 1171 | Officers in armed forces | police and protective services |
| 1172 | Senior police officers | police and protective services |
| 1173 | Senior officers in fire, ambulance, prison and related services | police and protective services |
| 1181 | Health services and public health managers and directors | healthcare-office based |
| 1184 | Social services managers and directors | social care |
| 1190 | Managers and directors in retail and wholesale | retail |
| 1211 | Managers and proprietors in agriculture and horticulture | other workers-non-office based |
| 1213 | Managers and proprietors in forestry, fishing and related services | other workers-non-office based |
| 1221 | Hotel and accommodation managers and proprietors | hospitality |
| 1223 | Restaurant and catering establishment managers and proprietors | hospitality |
| 1224 | Publicans and managers of licensed premises | hospitality |
| 1225 | Leisure and sports managers | other workers-office based |
| 1226 | Travel agency managers and proprietors | other workers-office based |
| 1241 | Health care practice managers | healthcare-office based |
| 1242 | Residential, day and domiciliary care managers and proprietors | social care |
| 1251 | Property, housing and estate managers | other workers-non-office based |
| 1252 | Garage managers and proprietors | other workers-non-office based |
| 1253 | Hairdressing and beauty salon managers and proprietors | personal care |
| 1254 | Shopkeepers and proprietors – wholesale and retail | retail |
| 1255 | Waste disposal and environmental services managers | sanitation services |
| 1259 | Managers and proprietors in other services n.e.c. | other workers-office based |
| 2111 | Chemical scientists | other workers-non-office based |
| 2112 | Biological scientists and biochemists | other workers-non-office based |
| 2113 | Physical scientists | other workers-non-office based |
| 2114 | Social and humanities scientists | other workers-non-office based |
| 2119 | Natural and social science professionals n.e.c. | other workers-office based |
| 2121 | Civil engineers | other workers-non-office based |
| 2122 | Mechanical engineers | other workers-non-office based |
| 2123 | Electrical engineers | other workers-non-office based |
| 2124 | Electronics engineers | other workers-non-office based |
| 2126 | Design and development engineers | other workers-non-office based |
| 2127 | Production and process engineers | other workers-non-office based |
| 2129 | Engineering professionals n.e.c. | other workers-non-office based |
| 2133 | IT specialist managers | other workers-office based |
| 2134 | IT project and programme managers | other workers-office based |
| 2135 | IT business analysts, architects and systems designers | other workers-office based |
| 2136 | Programmers and software development professionals | other workers-office based |
| 2137 | Web design and development professionals | other workers-office based |
| 2139 | Information technology and telecommunications professionals n.e.c. | other workers-office based |
| 2141 | Conservation professionals | other workers-non-office based |
| 2142 | Environment professionals | other workers-non-office based |
| 2150 | Air traffic controllers | other workers-office based |
| 2211 | Medical practitioners | healthcare-patient contact |
| 2212 | Psychologists | healthcare-patient contact |
| 2213 | Pharmacists | healthcare-patient contact |
| 2214 | Ophthalmic opticians | healthcare-patient contact |
| 2215 | Dental practitioners | healthcare-patient contact |
| 2216 | Veterinarians | other workers-non-office based |
| 2217 | Medical radiographers | healthcare-patient contact |
| 2218 | Podiatrists | healthcare-patient contact |
| 2219 | Health professionals n.e.c. | healthcare-patient contact |
| 2221 | Physiotherapists | healthcare-patient contact |
| 2222 | Occupational therapists | healthcare-patient contact |
| 2223 | Speech and language therapists | healthcare-patient contact |
| 2229 | Therapy professionals n.e.c. | healthcare-patient contact |
| 2231 | Nurses | healthcare-patient contact |
| 2232 | Midwives | healthcare-patient contact |
| 2311 | Higher education teaching professionals | education |
| 2312 | Further education teaching professionals | education |
| 2314 | Secondary education teaching professionals | education |
| 2315 | Primary and nursery education teaching professionals | education |
| 2316 | Special needs education teaching professionals | education |
| 2317 | Senior professionals of educational establishments | education |
| 2318 | Education advisers and school inspectors | education |
| 2319 | Teaching and other educational professionals n.e.c. | education |
| 2412 | Barristers and judges | other workers-office based |
| 2413 | Solicitors | other workers-office based |
| 2419 | Legal professionals n.e.c. | other workers-office based |
| 2421 | Chartered and certified accountants | other workers-office based |
| 2423 | Management consultants and business analysts | other workers-office based |
| 2424 | Business and financial project management professionals | other workers-office based |
| 2425 | Actuaries, economists and statisticians | other workers-office based |
| 2426 | Business and related research professionals | other workers-office based |
| 2429 | Business, research and administrative professionals n.e.c. | other workers-office based |
| 2431 | Architects | other workers-office based |
| 2432 | Town planning officers | other workers-office based |
| 2433 | Quantity surveyors | other workers-office based |
| 2434 | Chartered surveyors | other workers-office based |
| 2435 | Chartered architectural technologists | other workers-office based |
| 2436 | Construction project managers and related professionals | other workers-non-office based |
| 2442 | Social workers | social care |
| 2443 | Probation officers | social care |
| 2444 | Clergy | social care |
| 2449 | Welfare professionals n.e.c. | social care |
| 2451 | Librarians | other workers-office based |
| 2452 | Archivists and curators | other workers-office based |
| 2461 | Quality control and planning engineers | other workers-non-office based |
| 2462 | Quality assurance and regulatory professionals | other workers-office based |
| 2463 | Environmental health professionals | other workers-non-office based |
| 2471 | Journalists, newspaper and periodical editors | other workers-office based |
| 2472 | Public relations professionals | other workers-office based |
| 2473 | Advertising accounts managers and creative directors | other workers-office based |
| 3111 | Laboratory technicians | other workers-non-office based |
| 3112 | Electrical and electronics technicians | other workers-non-office based |
| 3113 | Engineering technicians | other workers-non-office based |
| 3114 | Building and civil engineering technicians | other workers-non-office based |
| 3115 | Quality assurance technicians | other workers-non-office based |
| 3116 | Planning, process and production technicians | other workers-non-office based |
| 3119 | Science, engineering and production technicians n.e.c. | other workers-non-office based |
| 3121 | Architectural and town planning technicians | other workers-non-office based |
| 3122 | Draughtspersons | other workers-non-office based |
| 3131 | IT operations technicians | other workers-office based |
| 3132 | IT user support technicians | other workers-office based |
| 3213 | Paramedics | healthcare-patient contact |
| 3216 | Dispensing opticians | healthcare-patient contact |
| 3217 | Pharmaceutical technicians | healthcare-patient contact |
| 3218 | Medical and dental technicians | healthcare-patient contact |
| 3219 | Health associate professionals n.e.c. | healthcare-patient contact |
| 3231 | Youth and community workers | social care |
| 3233 | Child and early years officers | social care |
| 3234 | Housing officers | social care |
| 3235 | Counsellors | social care |
| 3239 | Welfare and housing associate professionals n.e.c. | social care |
| 3311 | NCOs and other ranks | police and protective services |
| 3312 | Police officers (sergeant and below) | police and protective services |
| 3313 | Fire service officers (watch manager and below) | police and protective services |
| 3314 | Prison service officers (below principal officer) | police and protective services |
| 3315 | Police community support officers | police and protective services |
| 3319 | Protective service associate professionals n.e.c. | police and protective services |
| 3411 | Artists | other workers-office based |
| 3412 | Authors, writers and translators | other workers-office based |
| 3413 | Actors, entertainers and presenters | other workers-non-office based |
| 3414 | Dancers and choreographers | other workers-non-office based |
| 3415 | Musicians | other workers-non-office based |
| 3416 | Arts officers, producers and directors | other workers-non-office based |
| 3417 | Photographers, audio-visual and broadcasting equipment operators | other workers-non-office based |
| 3421 | Graphic designers | other workers-office based |
| 3422 | Product, clothing and related designers | other workers-office based |
| 3441 | Sports players | other workers-non-office based |
| 3442 | Sports coaches, instructors and officials | other workers-non-office based |
| 3443 | Fitness instructors | other workers-non-office based |
| 3511 | Air traffic controllers | other workers-office based |
| 3512 | Aircraft pilots and flight engineers | transport-public facing |
| 3513 | Ship and hovercraft officers | transport-nonpublic facing |
| 3520 | Legal associate professionals | other workers-office based |
| 3531 | Estimators, valuers and assessors | other workers-office based |
| 3532 | Brokers | other workers-office based |
| 3533 | Insurance underwriters | other workers-office based |
| 3534 | Finance and investment analysts and advisers | other workers-office based |
| 3535 | Taxation experts | other workers-office based |
| 3536 | Importers and exporters | other workers-office based |
| 3537 | Financial and accounting technicians | other workers-office based |
| 3538 | Financial accounts managers | other workers-office based |
| 3539 | Business and related associate professionals n.e.c. | other workers-office based |
| 3541 | Buyers and procurement officers | other workers-office based |
| 3542 | Business sales executives | other workers-office based |
| 3543 | Marketing associate professionals | other workers-office based |
| 3544 | Estate agents and auctioneers | other workers-office based |
| 3545 | Sales accounts and business development managers | other workers-office based |
| 3546 | Conference and exhibition managers and organisers | other workers-office based |
| 3550 | Conservation and environmental associate professionals | other workers-non-office based |
| 3561 | Public services associate professionals | other workers-office based |
| 3562 | Human resources and industrial relations officers | other workers-office based |
| 3563 | Vocational and industrial trainers and instructors | other workers-office based |
| 3564 | Careers advisers and vocational guidance specialists | other workers-office based |
| 3565 | Inspectors of standards and regulations | other workers-non-office based |
| 3567 | Health and safety officers | other workers-non-office based |
| 4112 | National government administrative occupations | other workers-office based |
| 4113 | Local government administrative occupations | other workers-office based |
| 4114 | Officers of non-governmental organisations | other workers-office based |
| 4121 | Credit controllers | other workers-office based |
| 4122 | Book-keepers, payroll managers and wages clerks | other workers-office based |
| 4123 | Bank and post office clerks | other workers-office based |
| 4124 | Finance officers | other workers-office based |
| 4129 | Financial administrative occupations n.e.c. | other workers-office based |
| 4131 | Records clerks and assistants | other workers-office based |
| 4132 | Pensions and insurance clerks and assistants | other workers-office based |
| 4133 | Stock control clerks and assistants | retail |
| 4134 | Transport and distribution clerks and assistants | transport-nonpublic facing |
| 4135 | Library clerks and assistants | other workers-office based |
| 4138 | Human resources administrative occupations | other workers-office based |
| 4151 | Sales administrators | retail |
| 4159 | Other administrative occupations n.e.c. | other workers-office based |
| 4161 | Office managers | other workers-office based |
| 4162 | Office supervisors | other workers-office based |
| 4211 | Medical secretaries | healthcare-office based |
| 4212 | Legal secretaries | other workers-office based |
| 4213 | School secretaries | education |
| 4214 | Company secretaries | other workers-office based |
| 4215 | Personal assistants and other secretaries | other workers-office based |
| 4216 | Receptionists | other workers-office based |
| 4217 | Typists and related keyboard occupations | other workers-office based |
| 5111 | Farmers | manual |
| 5112 | Horticultural trades | manual |
| 5113 | Gardeners and landscape gardeners | manual |
| 5114 | Groundsmen and greenkeepers | manual |
| 5119 | Agricultural and fishing trades n.e.c. | manual |
| 5211 | Smiths and forge workers | manual |
| 5212 | Moulders, core makers and die casters | manual |
| 5213 | Sheet metal workers | manual |
| 5214 | Metal plate workers, and riveters | manual |
| 5215 | Welding trades | manual |
| 5216 | Pipe fitters | manual |
| 5221 | Metal machining setters and setter-operators | manual |
| 5222 | Tool makers, tool fitters and markers-out | manual |
| 5223 | Metal working production and maintenance fitters | manual |
| 5224 | Precision instrument makers and repairers | manual |
| 5225 | Air-conditioning and refrigeration engineers | manual |
| 5231 | Vehicle technicians, mechanics and electricians | manual |
| 5232 | Vehicle body builders and repairers | manual |
| 5234 | Vehicle paint technicians | manual |
| 5235 | Aircraft maintenance and related trades | manual |
| 5236 | Boat and ship builders and repairers | manual |
| 5237 | Rail and rolling stock builders and repairers | manual |
| 5241 | Electricians and electrical fitters | other workers-non-office based |
| 5242 | Telecommunications engineers | other workers-non-office based |
| 5244 | TV, video and audio engineers | other workers-non-office based |
| 5245 | IT engineers | other workers-non-office based |
| 5249 | Electrical and electronic trades n.e.c. | other workers-non-office based |
| 5250 | Skilled metal, electrical and electronic trades supervisors | manual |
| 5311 | Steel erectors | manual |
| 5312 | Bricklayers and masons | manual |
| 5313 | Roofers, roof tilers and slaters | manual |
| 5314 | Plumbers and heating and ventilating engineers | manual |
| 5315 | Carpenters and joiners | manual |
| 5316 | Glaziers, window fabricators and fitters | manual |
| 5319 | Construction and building trades n.e.c. | manual |
| 5321 | Plasterers | manual |
| 5322 | Floorers and wall tilers | manual |
| 5323 | Painters and decorators | manual |
| 5330 | Construction and building trades supervisors | manual |
| 5411 | Weavers and knitters | manual |
| 5412 | Upholsterers | manual |
| 5413 | Footwear and leather working trades | manual |
| 5414 | Tailors and dressmakers | manual |
| 5419 | Textiles, garments and related trades n.e.c. | manual |
| 5421 | Pre-press technicians | manual |
| 5422 | Printers | manual |
| 5423 | Print finishing and binding workers | manual |
| 5431 | Butchers | food processing |
| 5432 | Bakers and flour confectioners | food processing |
| 5433 | Fishmongers and poultry dressers | food processing |
| 5434 | Chefs | hospitality |
| 5435 | Cooks | hospitality |
| 5436 | Catering and bar managers | hospitality |
| 5441 | Glass and ceramics makers, decorators and finishers | manual |
| 5442 | Furniture makers and other craft woodworkers | manual |
| 5443 | Florists | retail |
| 5449 | Other skilled trades n.e.c. | manual |
| 6121 | Nursery nurses and assistants | education |
| 6122 | Childminders and related occupations | education |
| 6123 | Playworkers | education |
| 6125 | Teaching assistants | education |
| 6126 | Educational support assistants | education |
| 6131 | Veterinary nurses | other workers-non-office based |
| 6132 | Pest control officers | sanitation services |
| 6139 | Animal care services occupations n.e.c. | other workers-non-office based |
| 6141 | Nursing auxiliaries and assistants | healthcare-patient contact |
| 6142 | Ambulance staff (excluding paramedics) | healthcare-patient contact |
| 6143 | Dental nurses | healthcare-patient contact |
| 6144 | Houseparents and residential wardens | social care |
| 6145 | Care workers and home carers | social care |
| 6146 | Senior care workers | social care |
| 6147 | Care escorts | social care |
| 6148 | Undertakers, mortuary and crematorium assistants | social care |
| 6211 | Sports and leisure assistants | other workers-non-office based |
| 6212 | Travel agents | other workers-office based |
| 6214 | Air travel assistants | transport-public facing |
| 6215 | Rail travel assistants | transport-public facing |
| 6219 | Leisure and travel service occupations n.e.c. | transport-public facing |
| 6221 | Hairdressers and barbers | personal care |
| 6222 | Beauticians and related occupations | personal care |
| 6231 | Housekeepers and related occupations | hospitality |
| 6232 | Caretakers | hospitality |
| 6240 | Cleaning and housekeeping managers and supervisors | hospitality |
| 7111 | Sales and retail assistants | retail |
| 7112 | Retail cashiers and check-out operators | retail |
| 7113 | Telephone salespersons | other workers-office based |
| 7114 | Pharmacy and other dispensing assistants | healthcare-patient contact |
| 7115 | Vehicle and parts salespersons and advisers | retail |
| 7121 | Collector salespersons and credit agents | other workers-office based |
| 7122 | Debt, rent and other cash collectors | other workers-office based |
| 7123 | Roundspersons and van salespersons | other workers-non-office based |
| 7124 | Market and street traders and assistants | retail |
| 7125 | Merchandisers and window dressers | other workers-non-office based |
| 7129 | Sales related occupations n.e.c. | retail |
| 7130 | Sales supervisors | retail |
| 7211 | Call and contact centre occupations | other workers-office based |
| 7213 | Telephonists | other workers-office based |
| 7214 | Communication operators | other workers-office based |
| 7215 | Market research interviewers | other workers-office based |
| 7219 | Customer service occupations n.e.c. | other workers-office based |
| 7220 | Customer service managers and supervisors | other workers-office based |
| 8111 | Food, drink and tobacco process operatives | food processing |
| 8112 | Glass and ceramics process operatives | manual |
| 8113 | Textile process operatives | manual |
| 8114 | Chemical and related process operatives | manual |
| 8115 | Rubber process operatives | manual |
| 8116 | Plastics process operatives | manual |
| 8117 | Metal making and treating process operatives | manual |
| 8118 | Electroplaters | manual |
| 8119 | Process operatives n.e.c. | manual |
| 8121 | Paper and wood machine operatives | manual |
| 8122 | Coal mine operatives | manual |
| 8123 | Quarry workers and related operatives | manual |
| 8124 | Energy plant operatives | manual |
| 8125 | Metal working machine operatives | manual |
| 8126 | Water and sewerage plant operatives | manual |
| 8127 | Printing machine assistants | manual |
| 8129 | Plant and machine operatives n.e.c. | manual |
| 8131 | Assemblers (electrical and electronic products) | manual |
| 8132 | Assemblers (vehicles and metal goods) | manual |
| 8133 | Routine inspectors and testers | manual |
| 8134 | Weighers, graders and sorters | manual |
| 8135 | Tyre, exhaust and windscreen fitters | manual |
| 8137 | Sewing machinists | manual |
| 8139 | Assemblers and routine operatives n.e.c. | manual |
| 8141 | Scaffolders, stagers and riggers | manual |
| 8142 | Road construction operatives | manual |
| 8143 | Rail construction and maintenance operatives | manual |
| 8149 | Construction operatives n.e.c. | manual |
| 8211 | Large goods vehicle drivers | transport-nonpublic facing |
| 8212 | Van drivers | transport-nonpublic facing |
| 8213 | Bus and coach drivers | transport-public facing |
| 8214 | Taxi and cab drivers and chauffeurs | transport-public facing |
| 8215 | Driving instructors | transport-public facing |
| 8221 | Crane drivers | other workers-non-office based |
| 8222 | Fork-lift truck drivers | other workers-non-office based |
| 8223 | Agricultural machinery drivers | other workers-non-office based |
| 8229 | Mobile machine drivers and operatives n.e.c. | other workers-non-office based |
| 8231 | Train and tram drivers | transport-nonpublic facing |
| 8232 | Marine and waterways transport operatives | transport-nonpublic facing |
| 8233 | Air transport operatives | transport-nonpublic facing |
| 8234 | Rail transport operatives | transport-nonpublic facing |
| 8239 | Other drivers and transport operatives n.e.c. | transport-nonpublic facing |
| 9111 | Farm workers | manual |
| 9112 | Forestry workers | manual |
| 9119 | Fishing and other elementary agriculture occupations n.e.c. | manual |
| 9120 | Elementary construction occupations | manual |
| 9132 | Industrial cleaning process occupations | sanitation services |
| 9134 | Packers, bottlers, canners and fillers | manual |
| 9139 | Elementary process plant occupations n.e.c. | manual |
| 9211 | Postal workers, mail sorters, messengers and couriers | transport-nonpublic facing |
| 9219 | Elementary administration occupations n.e.c. | other workers-office based |
| 9231 | Window cleaners | sanitation services |
| 9232 | Street cleaners | sanitation services |
| 9233 | Cleaners and domestics | sanitation services |
| 9234 | Launderers, dry cleaners and pressers | sanitation services |
| 9235 | Refuse and salvage occupations | sanitation services |
| 9236 | Vehicle valeters and cleaners | sanitation services |
| 9239 | Elementary cleaning occupations n.e.c. | sanitation services |
| 9241 | Security guards and related occupations | police and protective services |
| 9242 | Parking and civil enforcement occupations | police and protective services |
| 9244 | School midday and crossing patrol occupations | other workers-non-office based |
| 9249 | Elementary security occupations n.e.c. | police and protective services |
| 9251 | Shelf fillers | retail |
| 9259 | Elementary sales occupations n.e.c. | retail |
| 9260 | Elementary storage occupations | manual |
| 9271 | Hospital porters | healthcare-patient contact |
| 9272 | Kitchen and catering assistants | food processing |
| 9273 | Waiters and waitresses | hospitality |
| 9274 | Bar staff | hospitality |
| 9275 | Leisure and theme park attendants | hospitality |
| 9279 | Other elementary services occupations n.e.c. | hospitality |

**Supplementary Table 1: Occupational groupings used in the analysis.**

| Occupation | Moderna | Oxford/AstraZenica | Pfizer/BioNTech | Other | Missing |
| --- | --- | --- | --- | --- | --- |
| Education | 329 | 7,827 | 3,709 | * | * |
| % | 3 | 66 | 31 | * | * |
| Food processing | 40 | 618 | 383 | * | * |
| % | 3.84 | 59.25 | 36.72 | * | * |
| Healthcare-office based | * | 251 | 511 | * | * |
| % | * | 33 | 66 | * | * |
| Healthcare-patient contact | 54 | 2,787 | 7,950 | 28 | 27 |
| % | 0.5 | 26 | 73 | 0.26 | 0.25 |
| Hospitality | 91 | 1,354 | 932 | * | * |
| % | 4 | 57 | 39 | * | * |
| Manual | 329 | 6,311 | 3,379 | * | * |
| % | 3 | 63 | 34 | * | * |
| Other workers-non-office-based | 449 | 6,158 | 4,417 | 31 | 10 |
| % | 4 | 56 | 40 | 0.28 | 0.09 |
| Other workers-office-based | 3,028 | 39,654 | 25,953 | 170 | 63 |
| % | 4.4 | 57.58 | 37.69 | 0.25 | 0.09 |
| Personal care | 22 | 408 | 267 | * | * |
| % | 3.14 | 58 | 38 | * | * |
| Police and protective | 81 | 1,975 | 952 | * | * |
| % | 2.69 | 66 | 32 | * | * |
| Retail | 182 | 3,346 | 2,154 | * | * |
| % | 3.19 | 59 | 38 | * | * |
| Sanitation services | 29 | 1,030 | 493 | * | * |
| % | 1.86 | 66 | 32 | * | * |
| Social care | 70 | 2,949 | 3,362 | * | * |
| % | 1.1 | 46 | 53 | * | * |
| Transport-nonpublic facing | 77 | 1,946 | 846 | * | * |
| % | 2.68 | 68 | 29 | * | * |
| Transport-public facing | 22 | 728 | 313 | * | * |
| % | 2 | 68 | 29 | * | * |
| Not working/ student | 1,417 | 39,087 | 22,170 | 140 | 77 |
| % | 2 | 62 | 35 | 0.22 | 0.12 |
| Missing | * | 27,963 | 19,026 | 118 | * |
| % | * | 57 | 39 | 0.24 | * |

**Supplementary Table 2: Vaccine type for those receiving a first dose, by occupational group. Due to disclosure rules, counts below 10 are redacted. Values are also redacted where revealing the value would indirectly disclose a count below 10 by subtraction.**

| Occupation | Moderna | Oxford/AstraZenica | Pfizer/BioNTech | Other | Missing |
| --- | --- | --- | --- | --- | --- |
| Education | 325 | 7,798 | 3,679 | 11 | 11 |
| % | 3 | 66 | 31 | 0.09 | 0.09 |
| Food processing | 39 | 614 | 374 | * | * |
| % | 3.79 | 59.73 | 36.38 | * | * |
| Healthcare-office based | * | 249 | 509 | * | * |
| % | * | 33 | 66 | * | * |
| Healthcare-patient contact | 53 | 2,756 | 7,905 | 22 | 35 |
| % | 0.49 | 26 | 73 | 0.2 | 0.32 |
| Hospitality | 85 | 1,350 | 913 | * | * |
| % | 4 | 57 | 39 | * | * |
| Manual | 324 | 6,283 | 3,292 | * | * |
| % | 3 | 63 | 33 | * | * |
| Other workers-non-office-based | 441 | 6,135 | 4,356 | 24 | 14 |
| % | 4 | 56 | 40 | 0.22 | 0.13 |
| Other workers-office-based | 2,984 | 39,543 | 25,648 | 110 | 124 |
| % | 4.36 | 57.8 | 37.49 | 0.16 | 0.18 |
| Personal care | 22 | 404 | 262 | * | * |
| % | 3.18 | 58 | 38 | * | * |
| Police and protective | 76 | 1,965 | 943 | * | * |
| % | 2.54 | 66 | 32 | * | * |
| Retail | 180 | 3,327 | 2,118 | * | * |
| % | 3.19 | 59 | 38 | * | * |
| Sanitation services | 30 | 1,023 | 482 | * | * |
| % | 1.95 | 66 | 31 | * | * |
| Social care | 69 | 2,931 | 3,334 | * | * |
| % | 1.09 | 46 | 53 | * | * |
| Transport-nonpublic facing | 75 | 1,938 | 832 | * | * |
| % | 2.63 | 68 | 29 | * | * |
| Transport-public facing | 19 | 726 | 308 | * | * |
| % | 2 | 69 | 29 | * | * |
| Not working/ student | 1,372 | 38,738 | 21,671 | 97 | 139 |
| % | 2 | 62 | 35 | 0.16 | 0.22 |
| Missing | * | 27,848 | 18,733 | 81 | * |
| % | * | 57 | 39 | 0.17 | * |

**Supplementary Table 3: Vaccine type for those receiving a second dose, by occupational group. Due to disclosure rules, values are redacted when counts are below 10. Values are also redacted where revealing the value would indirectly disclose a count below 10 by subtraction.**

| **Occupation** | Moderna | Oxford/AstraZenica | Pfizer/BioNTech | Other |
| --- | --- | --- | --- | --- |
| Education | 3,460 | * | 7,528 | * |
| % | 31 | * | 68 | * |
| Food processing | 260 | * | 600 | * |
| % | 30.23 | * | 69.77 | * |
| Healthcare-office based | 82 | * | 658 | * |
| % | 11.08 | * | 89 | * |
| Healthcare-patient contact | 829 | * | 9,225 | * |
| % | 8.23 | * | 92 | * |
| Hospitality | 597 | * | 1,418 | * |
| % | 30 | * | 70 | * |
| Manual | 2,833 | * | 5,812 | * |
| % | 33 | * | 67 | * |
| Other workers-non-office-based | 3,283 | * | 6,672 | * |
| % | 33 | * | 67 | * |
| Other workers-office-based | 20,677 | 51 | 42,692 | 15 |
| % | 32.6 | 0.08 | 67.3 | 0.02 |
| Personal care | 176 | * | 409 | * |
| % | 30.03 | * | 70 | * |
| Police and protective | 912 | * | 1,776 | * |
| % | 33.9 | * | 66 | * |
| Retail | 1,575 | * | 3,380 | * |
| % | 31.74 | * | 68 | * |
| Sanitation services | 380 | * | 987 | * |
| % | 27.78 | * | 72 | * |
| Social care | 1,025 | * | 4,767 | * |
| % | 17.68 | * | 82 | * |
| Transport-nonpublic facing | 846 | * | 1,697 | * |
| % | 33.27 | * | 67 | * |
| Transport-public facing | 274 | * | 672 | * |
| % | 29 | * | 71 | * |
| Not working/ student | 15,073 | 104 | 41,211 | 18 |
| % | 27 | 0 | 73 | 0.03 |
| Missing | 13,708 | 45 | 30,710 | 15 |
| % | 31 | 0 | 69 | 0.03 |

**Supplementary Table 4: Vaccine type for those receiving a third dose, by occupational group. Due to disclosure rules, values are redacted when counts are below 10. Values are also redacted where revealing the value would indirectly disclose a count below 10 by subtraction.**

| Number of vaccines received | Hazard Ratio  (95% CI) |
| --- | --- |
| 0 | Ref |
| 1 | 0.80 (0.76 to 0.85) |
| 2 | 0.71 (0.68 to 0.75) |
| 3 | 0.40 (0.38 to 0.41) |

**Supplementary Table 5: Hazard ratios (95% CIs) for number of vaccines received in relation to infection with SARS-CoV-2. Based on n = 256,598 individuals. Adjusted for occupation, age, sex, ethnicity, deprivation, region, urban or rural area, household size, and presence of pre-existing health conditions.**

|  | **Number of infections** | | | |
| --- | --- | --- | --- | --- |
| **Occupation** | **0** | **1** | **2** | **3** |
| Education | 8,846 | 3,209 | 76 | 0 |
|  | 72.92 | 26.45 | 0.63 | 0 |
| Food processing | 830 | 246 | * | * |
|  | 76.85 | 22.78 | * | * |
| Healthcare-office based | 612 | 162 | * | * |
|  | 78.76 | 20.85 | * | * |
| Healthcare-patient contact | 8,544 | 2,379 | 47 | 0 |
|  | 77.89 | 21.69 | 0.43 | 0 |
| Hospitality | 1,919 | 528 | 15 | 0 |
|  | 77.94 | 21.45 | 0.61 | 0 |
| Manual | 8,080 | 2,265 | 42 | 0 |
|  | 77.79 | 21.81 | 0.4 | 0 |
| Other workers-non-office-based | 8,892 | 2,403 | 56 | 0 |
|  | 78.34 | 21.17 | 0.49 | 0 |
| Other workers-office-based | 54,857 | 15,027 | * | * |
|  | 78.13 | 21.4 | * | * |
| Personal care | 539 | 189 | * | * |
|  | 73.63 | 25.82 | * | * |
| Police and protective | 2,327 | 733 | 16 | 0 |
|  | 75.65 | 23.83 | 0.52 | 0 |
| Retail | 4,598 | 1,257 | 20 | 0 |
|  | 78.26 | 21.4 | 0.34 | 0 |
| Sanitation services | 1,245 | 357 | * | * |
|  | 77.33 | 22.17 | * | * |
| Social care | 5,007 | 1,481 | 45 | 0 |
|  | 76.64 | 22.67 | 0.69 | 0 |
| Transport-nonpublic facing | 2,318 | 655 | 17 | 0 |
|  | 77.53 | 21.91 | 0.57 | 0 |
| Transport-public facing | 833 | 246 | * | * |
|  | 76.56 | 22.61 | * | * |
| Not working/student | 52,214 | 12,749 | * | * |
|  | 80.04 | 19.54 | * | * |
| Missing/incomplete | 38,479 | 11,335 | 276 | 0 |
|  | 76.82 | 22.63 | 0.55 | 0 |

**Supplementary Table 6: Number of infections by occupational group. Due to disclosure rules, values are redacted when counts are below 10. Values are also redacted where revealing the value would indirectly disclose a count below 10 by subtraction.**

|  | Model 1 | Model 2 | Model 3 |
| --- | --- | --- | --- |
| Education | 1.272 [1.226,1.320] | 1.252 [1.207,1.299] | 1.250 [1.204,1.297] |
| Food processing | 1.049 [0.931,1.181] | 1.041 [0.924,1.173] | 0.984 [0.874,1.109] |
| Healthcare-office based | 0.901 [0.775,1.049] | 0.906 [0.778,1.054] | 0.950 [0.816,1.105] |
| Healthcare-patient contact | 1.016 [0.974,1.059] | 1.016 [0.974,1.060] | 1.071 [1.027,1.117] |
| Hospitality | 1.084 [1.006,1.168] | 1.081 [1.003,1.165] | 1.033 [0.959,1.114] |
| Manual | 0.993 [0.951,1.036] | 0.995 [0.953,1.039] | 0.947 [0.907,0.990] |
| Other workers-non-office based | 0.991 [0.952,1.032] | 0.994 [0.955,1.036] | 0.982 [0.943,1.023] |
| Other workers-office based | 1 [1,1] | 1 [1,1] | 1 [1,1] |
| Personal care | 1.155 [1.011,1.319] | 1.150 [1.006,1.313] | 1.076 [0.942,1.229] |
| Police and protective services | 1.101 [1.025,1.182] | 1.113 [1.036,1.196] | 1.098 [1.022,1.179] |
| Retail | 0.991 [0.937,1.048] | 0.992 [0.938,1.049] | 0.959 [0.906,1.014] |
| Sanitation services | 1.032 [0.934,1.141] | 1.031 [0.933,1.140] | 0.986 [0.892,1.091] |
| Social care | 1.104 [1.048,1.162] | 1.100 [1.045,1.158] | 1.111 [1.055,1.169] |
| Transport-nonpublic facing | 1.057 [0.980,1.139] | 1.058 [0.981,1.140] | 1.007 [0.934,1.086] |
| Transport-public facing | 1.121 [1.001,1.256] | 1.116 [0.996,1.250] | 1.082 [0.966,1.213] |
| Not working/student | 0.939 [0.916,0.962] | 0.939 [0.916,0.963] | 0.906 [0.884,0.929] |
| Missing/incomplete | 0.887 [0.866,0.909] | 0.886 [0.865,0.908] | 0.881 [0.860,0.903] |

**Supplementary Table 7: Hazard ratios (95% CIs) corresponding to occupational group in relation to infection with SARS-CoV-2. Based on n = 256,598 individuals. Model 1: Adjusted for age and sex. Model 2: Additionally adjusted for ethnicity, deprivation, region, urban or rural area, household size, and presence of pre-existing health conditions. Model 3: Additionally adjusted for number of vaccines received.**

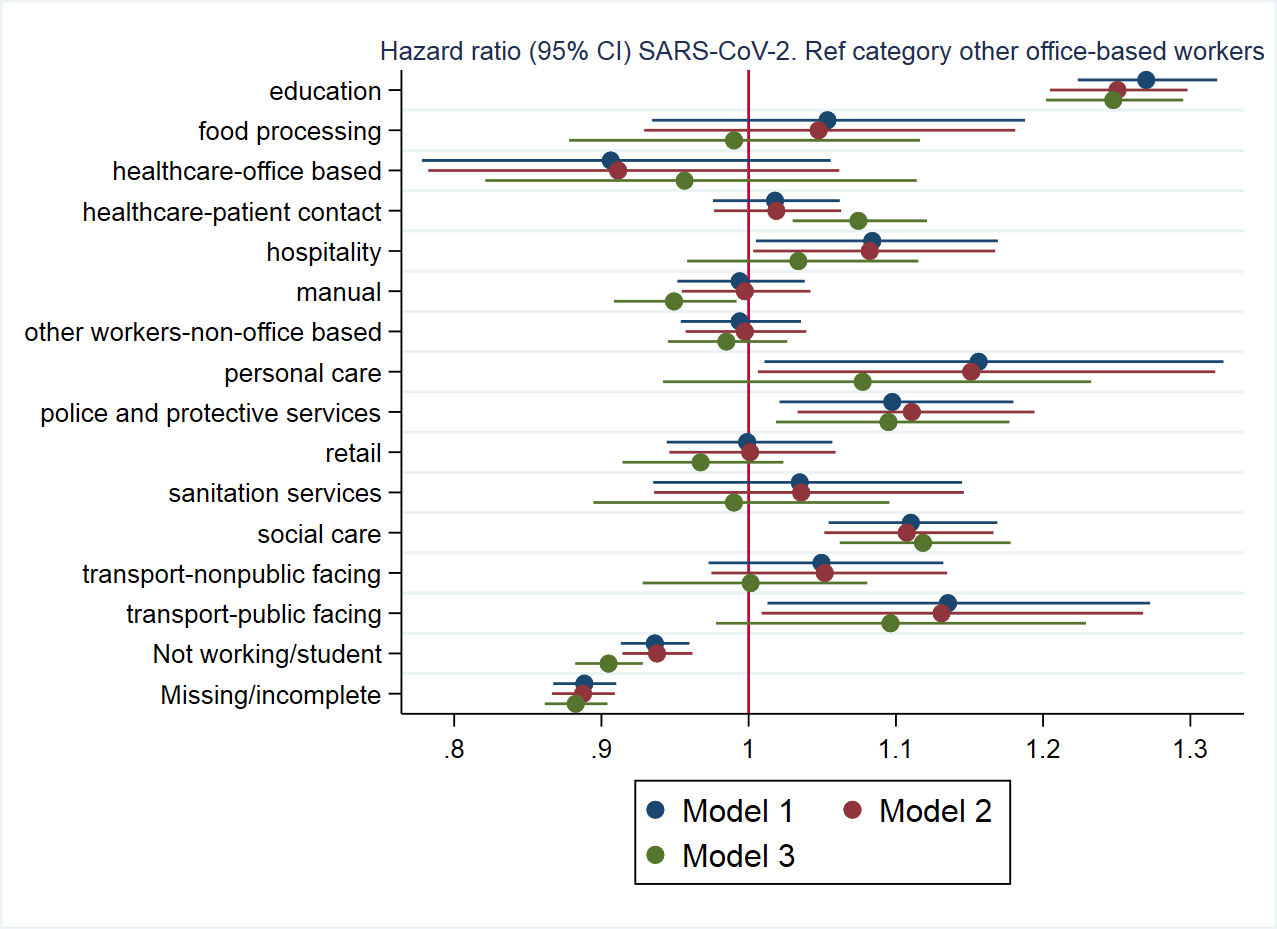

**Supplementary Figure 1: Sensitivity analysis, where a 6-month gap and negative PCR test is required between positive tests to register a new infection. Hazard ratios (95% CIs) corresponding to occupational group in relation to infection with SARS-CoV-2. Based on n = 256,598 individuals. Model 1: Adjusted for age and sex. Model 2: Additionally adjusted for ethnicity, deprivation, region, urban or rural area, household size, and presence of pre-existing health conditions. Model 3: Additionally adjusted for number of vaccines received.**

|  | Model 1 | Model 2 | Model 3 |
| --- | --- | --- | --- |
| education | 1.270 [1.224,1.318] | 1.250 [1.205,1.298] | 1.248 [1.202,1.295] |
| food processing | 1.054 [0.934,1.188] | 1.047 [0.929,1.181] | 0.990 [0.878,1.116] |
| healthcare-office based | 0.906 [0.778,1.056] | 0.911 [0.782,1.062] | 0.956 [0.821,1.114] |
| healthcare-patient contact | 1.018 [0.976,1.062] | 1.019 [0.977,1.063] | 1.075 [1.030,1.121] |
| hospitality | 1.084 [1.005,1.169] | 1.082 [1.003,1.168] | 1.034 [0.958,1.115] |
| manual | 0.994 [0.952,1.038] | 0.997 [0.955,1.042] | 0.949 [0.909,0.992] |
| other workers-non-office based | 0.994 [0.954,1.036] | 0.997 [0.957,1.039] | 0.985 [0.945,1.026] |
| other workers-office based | 1 [1,1] | 1 [1,1] | 1 [1,1] |
| personal care | 1.156 [1.011,1.323] | 1.151 [1.006,1.317] | 1.077 [0.942,1.233] |
| police and protective services | 1.098 [1.021,1.180] | 1.111 [1.033,1.194] | 1.095 [1.019,1.177] |
| retail | 0.999 [0.944,1.057] | 1.001 [0.946,1.059] | 0.967 [0.914,1.024] |
| sanitation services | 1.035 [0.935,1.145] | 1.036 [0.936,1.146] | 0.990 [0.895,1.096] |
| social care | 1.110 [1.054,1.169] | 1.107 [1.051,1.166] | 1.118 [1.062,1.178] |
| transport-nonpublic facing | 1.049 [0.973,1.132] | 1.052 [0.975,1.135] | 1.001 [0.928,1.081] |
| transport-public facing | 1.135 [1.013,1.273] | 1.131 [1.009,1.268] | 1.096 [0.978,1.229] |
| Not working/student | 0.936 [0.913,0.960] | 0.938 [0.914,0.962] | 0.905 [0.882,0.928] |
| Missing/incomplete | 0.888 [0.867,0.910] | 0.887 [0.866,0.909] | 0.883 [0.862,0.904] |

**Supplementary Table 8: Sensitivity analysis, where a 6-month gap and negative PCR test is required between positive tests to register a new infection. Hazard ratios (95% CIs) corresponding to occupational group in relation to infection with SARS-CoV-2. Based on n = 256,598 individuals. Model 1: Adjusted for age and sex. Model 2: Additionally adjusted for ethnicity, deprivation, region, urban or rural area, household size, and presence of pre-existing health conditions. Model 3: Additionally adjusted for number of vaccines received.**
